# Pathologic Nodal Upstaging Predictive Models for CT-based Clinical Node Negative Non-Small Cell Lung Cancer

**DOI:** 10.1101/2020.04.12.20063016

**Authors:** Weelic Chong, Yang Hai, Jian Zhou, Lun-xu Liu

## Abstract

**Background:** Accurate clinical nodal staging of non-small cell lung cancer (NSCLC) is essential for surgical management. Some clinical node negative cases diagnosed preoperatively by CT were later staged as pathological N1 (pN1) or pN2. Our study aimed to evaluate factors related to pathological nodal upstaging and develop statistical models for predicting upstaging.

**Methods:** We retrospectively reviewed 1,735 patients with clinical node negative NSCLC from 2011 to 2016 in the West China Lung Cancer database. Demographic and clinical data were analyzed via univariate and multivariate approaches. Predictive models were developed on a training set and validated with independent datasets.

**Results:** 171 (9.9%) clinical node negative patients have pathologic nodal upstaging to pN1. 191(11.0%) patients were upstaged to p(N1+N2). 91(5.2%) patients have pSN2 pathologic nodal upstaging. Preoperative factors were used to establish 3 statistical models for predicting pathological nodal upstaging. The area under the receiver operator characteristic (AUC) were 0.815, 0.768, and 0.726, for pN1, p(N1+N2) and pSN2 respectively.

**Conclusion:** Our models may help evaluate the possibility of nodal upstaging for clinical node negative NSCLC and enable surgeons to form appropriate plans preoperatively. External validation in a prospective multi-site study is needed before adoption into clinical practice.

## Introduction

Surgical resection is the primary and preferred treatment for suspected or confirmed Stage I and Stage II resectable non-small cell lung cancer (NSCLC) (1). Accurate Tumor Node Metastasis (TNM) staging is important for outcomes and treatment strategies(2). For example, N status is an important factor when surgeons make decisions on surgical methods, such as video-assisted thoracoscopic surgery (VATS) versus open thoracotomy or lobar versus limited resection, or stereotactic body radiotherapy (SBRT) for older patients(3–6). Meanwhile, studies have shown that neoadjuvant therapy improves the prognosis of patients with N2 stage IIIA disease(7,8).

To obtain more reliable clinical staging, there is an evolving array of procedures, each with distinct strengths – integrated positron emission tomography computed tomography (PET-CT), endobronchial ultrasound-guided transbronchial needle aspiration(EBUS-TBNA), esophageal ultrasound-guided fine-needle aspiration(EUS-FNA), and mediastinoscopy(9–14). The use of more specific testing needs to be balanced with the additional financial cost to the patient and the cost in terms of time that further testing takes away from treatment (definitive surgery). Thus, there is potential value in using upstaging prediction models based on easily obtainable clinical parameters.

Despite the importance of N staging in NSCLC, clinical and pathological nodal staging is discordant in a small proportion of patients(15–19). In a prospective trial, the discordance between clinical and pathological staging was investigated by PET/CT (20). Unsuspected N2 disease was found in 9% of patients who were clinically staged I, and 26% of patients clinically staged II. In centers that only routinely conduct low-dose CT, rates of unsuspected pathological upstaging may be higher.

Many prior models of NSCLC nodal upstaging utilize a mix of peri-operative and pre-operative data (16,19,21–23). In the operating room, it is not ideal for surgeons to wait for the peri-operative histology, use that data as input in a model, calculate the risk likelihood, and then make a snap decision in the operating room while performing critical procedures. Thus, models that utilize only pre-operative (without considering peri-operative variables such as the degree of tumor differentiation) is more useful to surgeons (Personal Communication, open peer review of other works(18)).

Thus, we only used clinical factors, and only selected clinical N0 NSCLC patient records for creating and validating our pathological upstaging models. We created 3 different statistical models for estimating the probability of nodal upstaging to pN1, p(N1+N2), and SN2 respectively.

## Material and Methods

### Patient population

A total of 4,336 patients who underwent surgical resection for primary lung cancer at West China Hospital, Sichuan University from January 2011 to December 2016 were reviewed retrospectively. All data were retrieved from Western China Lung Cancer database (WCLC). All patients were >18 years old with pathologically confirmed NSCLC after surgery. The eligibility criteria were: 1) clinical N0 NSCLC; 2) lobectomy or sublobar resection without microscopic residual tumor; 3) systematic lymph node dissection; 4) no previous history of other malignancies or received neoadjuvant chemotherapy or radiotherapy. 2,086 patients with cN0 NSCLC met our criteria above. We excluded patients with incomplete clinicopathological data, had no detailed records of preoperative examination, or had received chemotherapy/radiotherapy prior to surgery. Finally, 1,735 records were included for this study.

### Preoperative Evaluation

Preoperative assessment included physical examination, routine blood work, chest X-ray and computed tomography (CT), abdominal CT, brain magnetic resonance imaging (MRI), bone scintigraphy and/or bronchoscopy as necessary. CT was performed in all patients and interpreted independently by one radiologist and one surgeon. Surgery was conducted 1 to 2 weeks after CT was performed. Pre-operative clinical lung cancer staging was based on the 8th edition of TNM classification(25), with the modification that staging based on ground-glass opacity, i.e. staging as tumor in-situ or T1mi, was not conducted. Positive clinical nodal stage was defined by size >=1cm, or greater in the short-axis dimension. A central tumor was defined by a tumor visible by bronchoscopy while a peripheral tumor was defined by not visibxle. PET-CT is expensive and not covered by public medical insurance in China, so patients underwent PET-CT only if they are willing to afford the scan out-of-pocket.

### Pathology Evaluation

In the hospital, we routinely conduct an intraoperative frozen section (FS) to get an initial impression of the tumor and the lymph nodes. The intraoperative pathological findings were in the raw data, but not used for the univariate analysis or the predictive models.

Confirmatory postoperative histological classification was made with reference to the latest WHO guideline(24). The distribution of the positive nodes were based on final pathology reports of post-operative pathological nodal staging, as follows: (1) pN0 group; (2) pN1 (involving only N1 lymph nodes); (2) p(N1+N2) group (involving both N1 and N2 lymph nodes); and (3) pSN2 (skip nodal metastasis, in which there is direct metastasis to N2 station with no involvement of N1 lymph nodes).

### Statistical Analysis

Statistical analysis was performed using SPSS (Version 26.0, Chicago, IL, USA) and R 3.6.0 software (Institute for Statistics and Mathematics; www.r-project.org). Based on patient pathological nodal status, data was separated into 4 groups: pN0, pN1, p(N1+N2), and pSN2. Univariate analysis was conducted to identify clinicopathological characteristics that differ between the pN0 group and the upstaged groups. For univariate analysis of categorical variables, Fisher’s exact test (for N< 30) or χ2-test (for all other N) were used, while Student’s t-test was used for continuous variables. All P values were two-tailed with <0.05 considered to be statistically significant. For variable selection, a correlation matrix was created to visualize potential statistically significant differences. The identified independent variables were further manually selected for logistic regression. Before logistic regression was performed, the dataset was split into a training dataset and a test dataset in a 80:20 ratio. The training dataset was used for creating the logistic regression. The model was then tested in the testing dataset. Due to the unbalanced nature of the datasets (i.e. nodal upstaging is rare), accuracy was not used for determining the cut-off for the model. Instead, the optimal cut-off for the regression model was calculated using a cost function(26). For pN1 and p(N1+N2), the cost of a false negative (FN) was set to 100 and the cost of a false positive (FP) was set to 200. For pSN2, FN was set to 100 and FP was set to 600. The concordance between predicted and actually observed probabilities was examined with a confusion matrix. Nomograms for each model was created with rms 5.1 package in R(27). Area under the receiver operating characteristic curve (AUROC) analysis was performed to evaluate the discriminative capability of the model with pROC 1.16.1 (28). An exploratory validation using an external data set was also performed(18). Detailed statistical analyses are found in the supplementary text.

## Results

### Patient Characteristics

A total of 1,735 patients were enrolled, with 948(54.6%) males and 788(45.4%) females. The average age and standard deviation were 58.6 and 9.9, respectively. In our study, 1,053(60.7%) patients never smoked, 277(16.0%) patients had less than 30 pack-years, and 405(23.3%) patients had 30 or more pack-years. According to preoperative CT, 569(32.9%) tumors were located in the right upper lobe(RUL), 188(10.8%) tumors were in the right middle lobe(RML), and 299(17.2%) tumors were in the right lower lobe(RLL); 384(22.1%) tumors were in the left upper lobe(LUL), and 295(17.0%) tumors were in the left lower lobe(LLL). Centrally located tumors were found in 374(21.6%) cases by bronchoscopy. For clinical T stage based on CT findings, there were 107(6.2%) cases with T1a, 298(17.2%) cases with T1b, 222(12.8%) cases with T1c, 765(44.1%) cases with T2a, 177(10.2%) cases with T2b, 155(8.9%) cases with T3, and 11(0.6%) cases with T4. Visceral pleural involvement was found in 819(47.2%) cases, and tumors with solid components were identified in 1,284(74.1%) cases while pure ground-glass opacity (GGO) were found in 451(25.9%) cases. Video-assisted thoracic surgery (VATS) was performed in 1,300(75.0%) patients compared with thoracotomy in 435(25.0%) patients. Specimens were histologically proven to be lung adenocarcinoma in 1,345(77.5%) patients, squamous cell carcinoma in 345(19.9%) patients, or other NSCLC subtypes in 45(2.6%). As for the degree of differentiation, 22(1.3%) cases were well, 94(5.4%) cases were well-moderate, 1,135(65.4%) cases were moderate, 330(19.0%) were moderate-poor, and 154(8.9%) cases had poor differentiation. Details are presented in Table 1.

**Table 1.** Demographic Characteristics (N=1735)

| Variables | N (%) |
| --- | --- |
| Gender |  |
| Male | 948(54.6) |
| Female | 788(45.4) |
| Age (mean± SD) | 58.6± 9.9 |
| Smoking status (pack-years) |  |
| 0 (Never smoked) | 1053(60.7) |
| <30 | 277(16.0) |
| ≥30 | 405(23.3) |
| Location |  |
| RUL | 569(32.9) |
| RML | 188(10.8) |
| RLL | 299(17.2) |
| LUL | 384(22.1) |
| LLL | 295(17.0) |
| Central/Peripheral |  |
| Central | 374(21.6) |
| Peripheral | 1361(78.4) |
| T stage |  |
| T1a | 107(6.2) |
| T1b | 298(17.2) |
| T1c | 222(12.8) |
| T2a | 765(44.1) |
| T2b | 177(10.2) |
| T3 | 155(8.9) |
| T4 | 11(0.6) |
| Visceral pleura involvement |  |
| Yes | 819(47.2) |
| No | 916(52.8) |
| Consolidation |  |
| Solid | 1284(74.0) |
| GGO | 451(26.0) |
| Surgical procedure |  |
| VATS | 1300(75.0) |
| Thoracotomy | 435(25.0) |
| Histology |  |
| Adenocarcinoma | 1345(77.5) |
| Squamous cell carcinoma | 345(19.9) |
| Others | 45(2.6) |
| Differentiation |  |
| Well | 22(1.3) |
| Well-moderate | 94(5.4) |
| Moderate | 1135(65.4) |
| Moderate-poor | 330(19.0) |
| Poor | 154(8.9) |

NSCLC, non-small cell lung cancer; N(%), cases sample size and percentage; SD, standard deviation;
Brinkman index: the number of cigarettes smoked per day multiplied by the number of years of smoking
T stage, tumor; GGO, ground glass opaque; VATS, Video-assisted thoracoscopic surgery;
RUL, right upper lobe; RML, right medial lobe; RLL, right lower lobe; LUL, left upper lobe; LLL, left lower lobe

### Postoperative nodal status among patients with cN0 NSCLC

453(26.1%) cases were found to have positive nodes based on final pathology reports. 1282 patients (73.9%) were node negative. Of the 1735 cN0 cases, the rate of upstaging to pN1, p(N1+N2), and SN2 were 9.8%, 11.0%, and 5.2% Out of the nodal upstaging cases, 171(37.7%), 191(41.2%), and 91(20.1%) cases were diagnosed as pN1, p(N1+N2), and SN2, respectively.

### Risk Factors Predicting Nodal Upstaging to pN1

For the pN1 group, univariate analysis identified the following factors as significant predictors: male sex (P<0.001), smoking history (pack-years >=30) (P<0.001), tumor located in LLL (P<0.001), centrally located tumor (P<0.001), clinical T staging based on CT(P<0.001), visceral pleural involvement (P<0.001) and consolidation with solid component (P<0.001) as significant risk factors in univariate analysis (Table 2). Multivariate logistic regression identified male sex (odds ratio[OR],1.83;P<0.01), tumor located in LLL (OR,2.02;P=0.003), centrally located tumor (OR,2.35;P<0.001), visceral pleural involvement (OR,3.28;P<0.001), tumor with solid component (OR,3.89;P<0.001) and tumor size (OR,1.40;P<0.001) to be statistically significant (Table 4).

**Table 2.** Univariate Analysis for the Prevalence of pN1

| Variables | pN0 | pN1 | *P* value<sup>a</sup> |
| --- | --- | --- | --- |
|  | Number of patients (%) |  |  |
|  | 1282(88.2) | 171 (11.8) |  |
| Gender |  |  | **<0.001** |
| Male | 658(51.3) | 129(75.4) |  |
| Female | 624(48.7) | 42(24.6) |  |
| Age (mean± SD) | 58.8± 10.0 | 59.3± 8.4 | 0.549 |
| Smoking status (pack-years) |  |  | **<0.001** |
| 0 (Never smoked) | 817 (63.7) | 68 (39.8) |  |
| 0-30 | 201 (15.7) | 29 (17.0) |  |
| >=30 | 264 (20.6) | 74 (43.3) |  |
| Location |  |  | **<0.001** |
| RUL | 433(33.8) | 40(23.4) |  |
| RML | 140(10.9) | 23(13.5) |  |
| RLL | 226(17.6) | 24(14.0) |  |
| LUL | 280(21.8) | 34(19.9) |  |
| LLL | 203(15.8) | 50(29.2) |  |
| Central/Peripheral |  |  | **<0.001** |
| Central | 219(17.1) | 81(47.4) |  |
| Peripheral | 1063(82.9) | 90(52.6) |  |
| T stage |  |  | **<0.001** |
| T1a | 105(8.2) | 0(0.0) |  |
| T1b | 271(21.1) | 7(4.1) |  |
| T1c | 185(14.4) | 13(7.6) |  |
| T2a | 523(40.8) | 81(47.4) |  |
| T2b | 105(8.2) | 32(18.7) |  |
| T3 | 83(6.5) | 38(22.2) |  |
| T4 | 10(0.8) | 0(0.0) |  |
| Visceral pleura involvement |  |  | **<0.001** |
| Yes | 515(40.2) | 116(67.8) |  |
| No | 767(59.8) | 55(32.2) |  |
| Consolidation |  |  | **<0.001** |
| Solid | 872(68.0) | 157(91.8) |  |
| GGO | 410(32.0) | 14(8.2) |  |
| Histology |  |  | **<0.001** |
| Adenocarcinoma | 1019(79.5) | 99(57.9) |  |
| Squamous cell carcinoma | 235(18.3) | 64(37.4) |  |
| Others | 28(2.2) | 8(4.7) |  |

NSCLC, non-small cell lung cancer; N(%), cases sample size and percentage; SD, standard deviation;
N stage, node; T stage, tumor; GGO, ground glass opaque; RUL, right upper lobe; RML, right medial lobe;
RLL, right lower lobe; LUL, left upper lobe; LLL, left lower lobe
<sup>a</sup>Significance of Fisher's exact test, $\chi^2$ -test or t-test. *P*<0.05 was considered statistically significant.

**Table 3.** Univariate Analysis for the Prevalence of p(N1+N2) and SN2

| Variables | pN0 | pN1+N2 | *P* value <sup>a</sup> | pN0 | SN2 | *P* value <sup>a</sup> |
| --- | --- | --- | --- | --- | --- | --- |
|  | Number of patients (%) |  |  | Number of patients (%) |  |  |
|  | 1282(87.0) | 191(13.0) |  | 1282(93.4) | 91(6.6) |  |
| Gender |  |  | 0.106 |  |  | 0.384 |
| Male | 658(51.3) | 110(42.4) |  | 658(51.3) | 51(56.0) |  |
| Female | 624(48.7) | 81(57.6) |  | 624(48.7) | 40(44.0) |  |
| Age (mean±SD) | 58.8±10.0 | 56.4±9.8 | 0.823 | 58.8±10.0 | 56.9±10.1 | 0.956 |
| Smoking status (pack-years) |  |  | 0.09 |  |  | 0.617 |
| 0 (never smoked) | 817(63.7) | 108(56.5) |  | 817(63.7) | 60(65.9) |  |
| 0-30 | 201 (15.7) | 31 (16.2) |  | 201 (15.7) | 16 (17.6) |  |
| >= 30 | 264 (20.1) | 52 (27.2) |  | 264 (20.1) | 15 (16.5) |  |
| Location |  |  | 0.694 |  |  | 0.236 |
| RUL | 433(33.8) | 57(29.8) |  | 433(33.8) | 39(42.9) |  |
| RML | 140(10.9) | 19(9.9) |  | 140(10.9) | 6(6.6) |  |
| RLL | 226(17.6) | 33(17.3) |  | 226(17.6) | 16(17.6) |  |
| LUL | 280(21.8) | 49(25.7) |  | 280(21.8) | 21(23.1) |  |
| LLL | 203(15.8) | 33(17.3) |  | 203(15.8) | 9(9.9) |  |
| Central/Peripheral |  |  | <0.001 |  |  | 0.903 |
| Central | 219(17.1) | 58(30.4) |  | 219(17.1) | 16(17.6) |  |
| Peripheral | 1063(82.9) | 133(69.6) |  | 1063(82.9) | 75(82.4) |  |
| T stage |  |  | <0.001 |  |  | <0.001 |
| T1a | 105(8.2) | 2(1.0) |  | 105(12.0) | 0(0.0) |  |
| T1b | 271(21.1) | 12(6.3) |  | 271(32.8) | 8(8.8) |  |
| T1c | 185(14.4) | 13(6.8) |  | 185(25.4) | 11(12.1) |  |
| T2a | 523(40.8) | 103(53.9) |  | 523(14.4) | 58(63.7) |  |
| T2b | 105(8.2) | 33(17.3) |  | 105(8.2) | 7(7.7) |  |
| T3 | 83(6.5) | 27(14.1) |  | 83(6.5) | 7(7.7) |  |
| T4 | 10(0.8) | 1(0.5) |  | 10(0.8) | 0(0.0) |  |
| Visceral pleura involvement |  |  | **<0.001** |  |  | **<0.001** |
| Yes | 515(40.2) | 126(66.0) |  | 515(40.2) | 62(68.1) |  |
| No | 767(59.8) | 65(34.0) |  | 767(59.8) | 29(31.9) |  |
| Consolidation |  |  | **<0.001** |  |  | **<0.001** |
| Solid | 872(68.0) | 176(92.1) |  | 872(68.0) | 79(86.8) |  |
| GGO | 410(32.0) | 15(7.9) |  | 410(32.0) | 12(13.2) |  |
| Histology |  |  | 0.333 |  |  | 0.984 |
| Adenocarcinoma | 1019(79.5) | 154(80.6) |  | 1019(79.5) | 73(80.2) |  |
| Squamous cell carcinoma | 235(18.3) | 30(15.7) |  | 235(18.3) | 16(17.6) |  |
| Others | 28(2.2) | 7(3.7) |  | 28(2.2) | 2(2.2) |  |

NSCLC, non-small cell lung cancer; N(%), cases sample size and percentage; SD, standard deviation; Brinkman index: the number of cigarettes smoked per day multiplied by the number of years of smoking N stage, node; T stage, tumor; GGO, ground glass opaque; VATS, Video-assisted thoracoscopic surgery; RUL, right upper lobe; RML, right medial lobe; RLL, right lower lobe; LUL, left upper lobe; LLL, left lower lobe <sup>a</sup>Significance of Fisher's exact test, $\chi^2$ -test or t-test. P<0.05 was considered statistically significant.

**Table 4.**
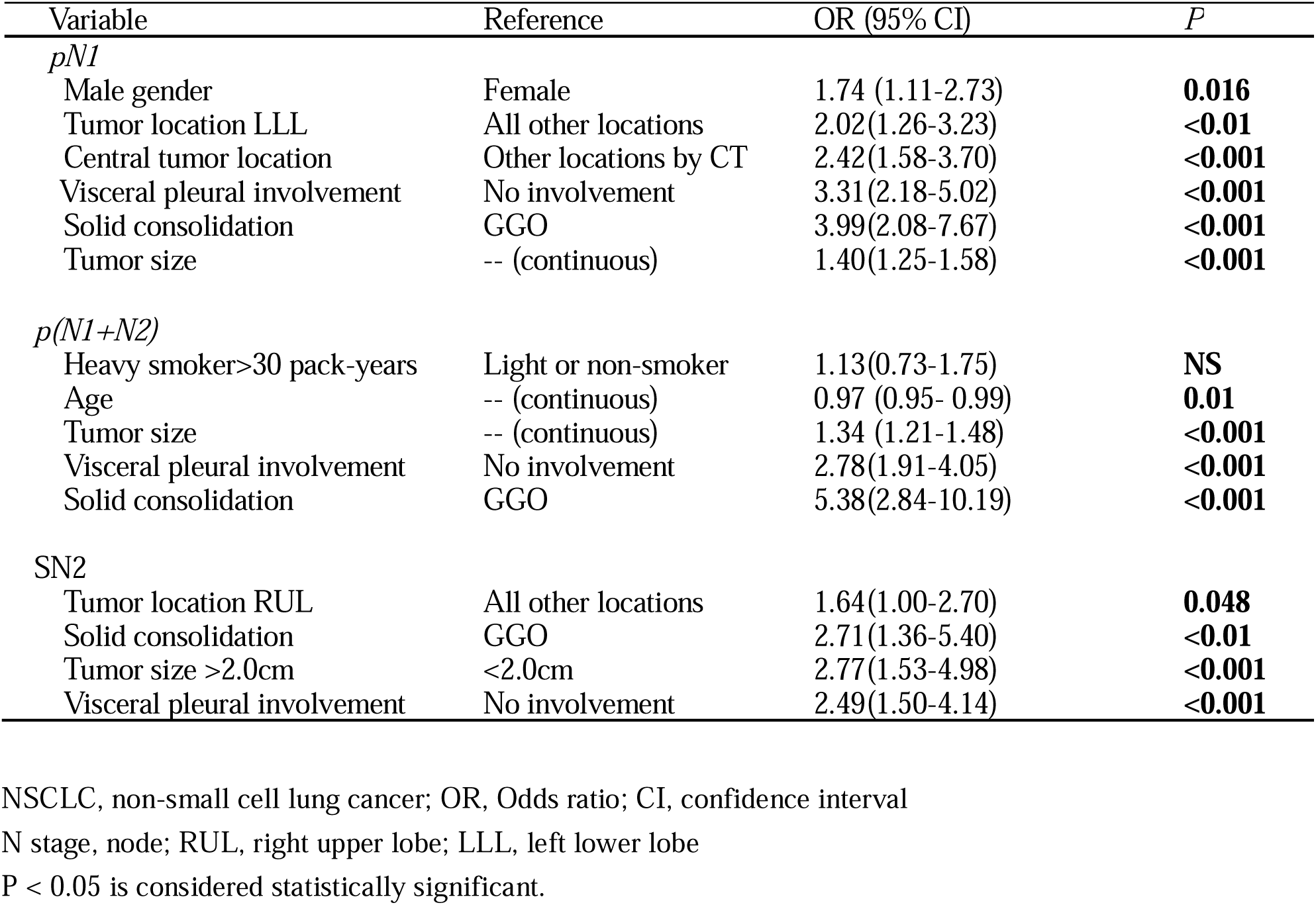
Multivariate Analysis for Prevalence of nodal upstaging

| Variable | Reference | OR (95% CI) | P |
| --- | --- | --- | --- |
| *pN1* |  |  |  |
| Male gender | Female | 1.74 (1.11-2.73) | **0.016** |
| Tumor location LLL | All other locations | 2.02(1.26-3.23) | **<0.01** |
| Central tumor location | Other locations by CT | 2.42(1.58-3.70) | **<0.001** |
| Visceral pleural involvement | No involvement | 3.31(2.18-5.02) | **<0.001** |
| Solid consolidation | GGO | 3.99(2.08-7.67) | **<0.001** |
| Tumor size | -- (continuous) | 1.40(1.25-1.58) | **<0.001** |
| *p(N1+N2)* |  |  |  |
| Heavy smoker>30 pack-years | Light or non-smoker | 1.13(0.73-1.75) | **NS** |
| Age | -- (continuous) | 0.97 (0.95- 0.99) | **0.01** |
| Tumor size | -- (continuous) | 1.34 (1.21-1.48) | **<0.001** |
| Visceral pleural involvement | No involvement | 2.78(1.91-4.05) | **<0.001** |
| Solid consolidation | GGO | 5.38(2.84-10.19) | **<0.001** |
| *SN2* |  |  |  |
| Tumor location RUL | All other locations | 1.64(1.00-2.70) | **0.048** |
| Solid consolidation | GGO | 2.71(1.36-5.40) | **<0.01** |
| Tumor size >2.0cm | <2.0cm | 2.77(1.53-4.98) | **<0.001** |
| Visceral pleural involvement | No involvement | 2.49(1.50-4.14) | **<0.001** |

NSCLC, non-small cell lung cancer; OR, Odds ratio; CI, confidence interval
N stage, node; RUL, right upper lobe; LLL, left lower lobe
P < 0.05 is considered statistically significant.

### Risk Factors Predicting Nodal Upstaging to p(N1+N2)

For the p(N1+N2) group, centrally located tumor (P<0.001), clinical T staging (P<0.001), visceral pleural involvement (P<0.001), tumors greater than 2.0cm (P<0.001), and consolidation with solid component (P<0.001) as significant risk factors in univariate analysis (Table 3). Multivariate logistic regression identified tumor size (OR,1.34;P<0.001), visceral pleural involvement (OR,2.78; P<0.001) and consolidation with solid component (OR, 5.38;P<0.001) to be statistically significant (Table 4).

### Risk Factors Predicting Nodal Upstaging to SN2

For the SN2 group, univariate analysis showed RUL-located tumors (P=0.041), clinical T stage (P<0.001), visceral pleural involvement (P<0.001), consolidation with solid component (P<0.001) as significant risk factors (Table 3). We also found tumors greater than 2.0 cm was a predictive risk factor (P<0.001). Using multivariate analysis, the following 4 factors were found with statistical significance: tumor located in RUL (OR, 1.64;P=0.048), tumor size>2.0cm (OR, 2.77;P<0.001), visceral pleural involvement (OR, 2.71; P<0.001) and consolidation with solid component (OR,2.71;P=0.005) (Table 4).

### Predictive models and validation

We identified pre-operative variables that were significant in the univariate test and in the correlation matrix (Figs 1a, 2a, 3a). From these, we only selected relevant pre-operative variables. We performed logistic regression on the selected variables and the binary nodal upstaging outcome to obtain three regression models: pN1 (Fig 1), p(N1+N2) (Fig 2), and pSN2 (Fig 3). The formulas are presented in Table 5. Internal validation was performed with the validation set, with similar performance obtained from the training and validation sets (Fig 1b, 2b, 3b). The cut-off was selected using a cost function that includes the cost of a false negative (i.e. patient with unsuspected upstaging), and the cost of a false positive (i.e. patient that has no upstaging, falsely predicted to have upstaging) (Figs 1c, 2c, 3c). The models’ predictions were compared to the actual result and displayed in a confusion matrix (Figs 1d, 2d, 3d). Static nomograms are presented in Figs 1 e, 2e, 3e. The AUC for the pN1 model was 0.815, p(N1+N2) was 0.768, and pSN2 was 0.726 (Figs 1f, 2f, 3f), indicating that all three models have good discriminatory ability, especially the pN1 model. We also conducted an exploratory external validation using data from an independent, previously published dataset from another institution, obtaining an AUC of 0.635 (Supplementary Fig 1).

**Table 5.** Summary of Logistic Regression Models

| Variable | Equation |
| --- | --- |
| *General equation for a binary logistic regression model* | $l = \log_e \left( \frac{\pi}{1 - \pi} \right) = \beta_0 + \beta_1 x_1 + \dots + \beta_{p-1} x_{p-1}$ |
| *pN1* | $\log \left( \frac{\pi}{1 - \pi} \right) = -5.68 + 0.70 * LLL + 0.86 * central$<br>$+ 0.33 * size + 1.19 * visceral + 1.36 * consolidation$<br>$+ 0.60 * gender$ |
| *p(N1+N2)* | $\log \left( \frac{\pi}{1 - \pi} \right) = -4.76 + 0.30 * size$<br>$0.99 * visceral + 1.69 * consolidation$ |
| *pSN2* | $\log \left( \frac{\pi}{1 - \pi} \right) = -4.80 + 0.50 * RUL + 1.02 * T2plus$<br>$+ 0.91 * visceral + 1.00 * consolidation$ |

List of variable abbreviations:
central: presence of tumor by bronchoscopy (1), absence (0)
size: size of tumor from CT (in cm). continuous variable
visceral: visceral pleural involvement identified by CT (1), absence (0)
consolidation: solid component identified by CT (1), ground-glass opacity on CT (0)
gender: male (1), female (0)
RUL: tumor located in right-upper lobe on CT (1), other locations (0)
T2plus: Tumor size is greater than 2.0cm (1), else equal to or less than 2.0 cm (0)

**Figure 1:**
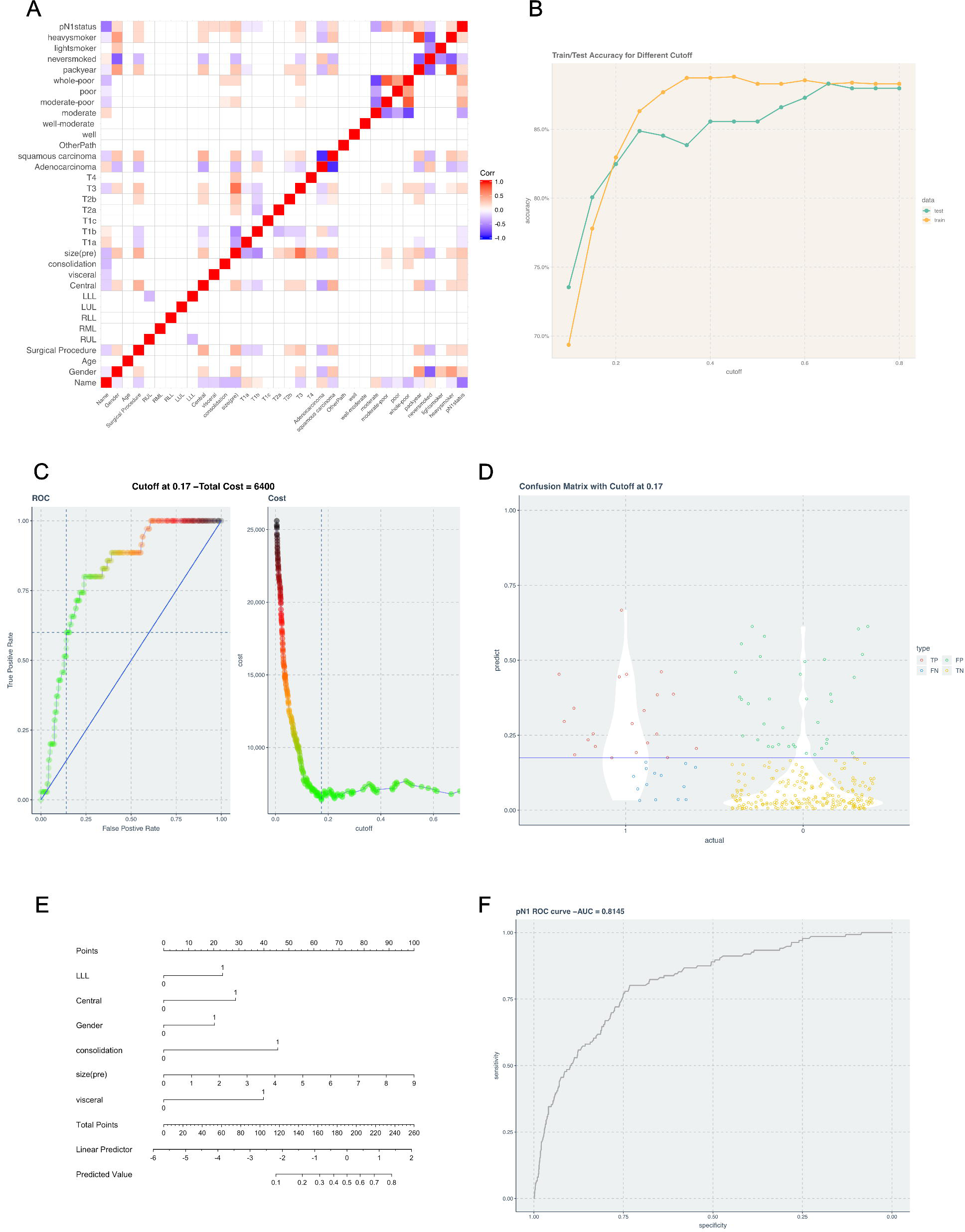
pN1 upstaging model from 1282 pN0 and 171 pN1 cases. **(a)** Correlation matrix identifies variables that are associated with upstaging (b) Accuracy measurements for training set and test set at different cutoffs. Accuracy does not have defined peak, showing it cannot be used to determine cutoff values in unbalanced datasets (c) Application of a cost function for determining the cutoff value. 0.17 is the optimal cutoff identified. (d) Confusion matrix at the cutoff in 1(c), symbolized by the horizontal line in blue. Points on the upper left are true positives, bottom left are false negatives, upper right are false positives, and bottom right are true negatives. (d) nomogram of the upstaging model (e) ROC (Receptor Operating Characteristic) curve. Area under the curve is 0.8145.

**Figure 2:**
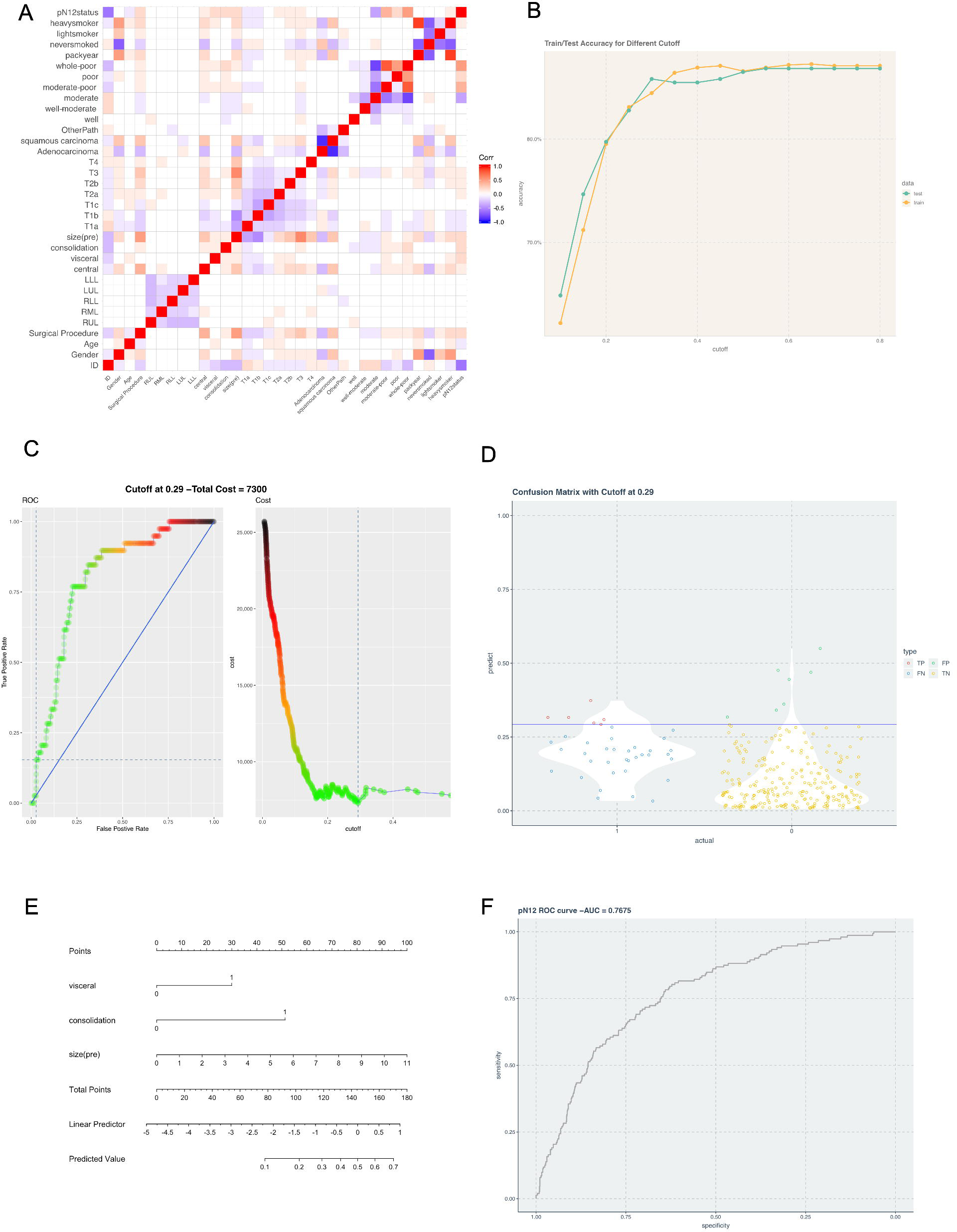
p(N1+N2) upstaging model from 1282 pN0 and 191 p(N1+N2) cases. **(a)** Correlation matrix identifies variables that are associated with upstaging (b) Accuracy measurements for training set and test set at different cutoffs. Accuracy does not have defined peak, showing it cannot be used to determine cutoff values in unbalanced datasets (c) Application of a cost function for determining the cutoff value. 0.29 is the optimal cutoff identified. (d) Confusion matrix at the cutoff in 2(c), symbolized by the horizontal line in blue. Points on the upper left are true positives, bottom left are false negatives, upper right are false positives, and bottom right are true negatives. (d) nomogram of the upstaging model (e) ROC (Receptor Operating Characteristic) curve. Area under the curve is 0.7675.

**Figure 3:**
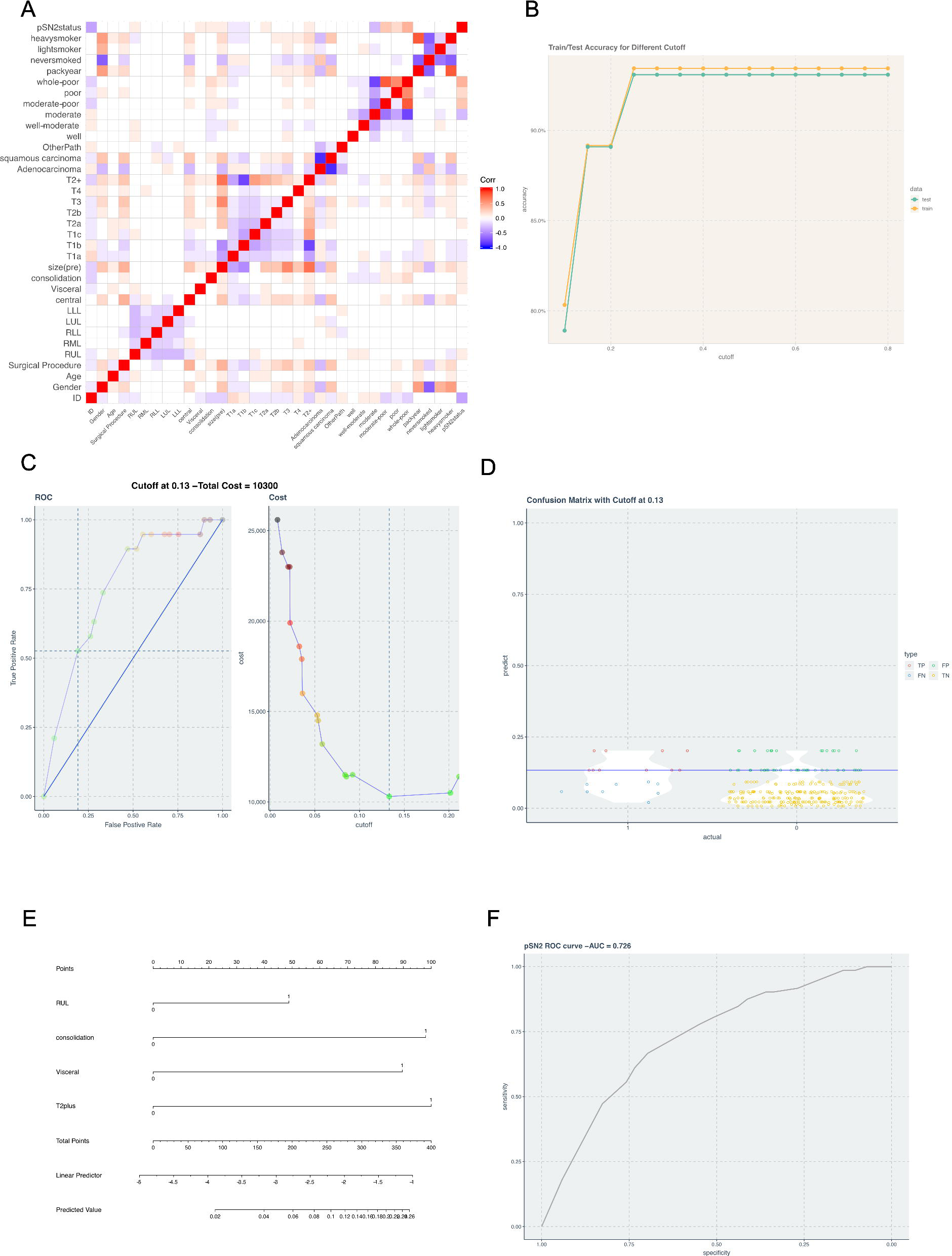
p(N1+N2) upstaging model from 1282 pN0 and 91 pSN2 cases. **(a)** Correlation matrix identifies variables that are associated with upstaging (b) Accuracy measurements for training set and test set at different cutoffs. Accuracy does not have defined peak, showing it cannot be used to determine cutoff values in unbalanced datasets (c) Application of a cost function for determining the cutoff value. 0.13 is the optimal cutoff identified. (d) Confusion matrix at the cutoff in 3(c), symbolized by the horizontal line in blue. Points on the upper left are true positives, bottom left are false negatives, upper right are false positives, and bottom right are true negatives. (d) nomogram of the upstaging model (e) ROC (Receptor Operating Characteristic) curve. Area under the curve is 0.726.

## Discussion

There are many reports of discordance between clinical and pathological staging for early-stage NSCLC. Bott et al. reported 9,530/55,653(17%) patients with more advanced staging on final pathologic review(15). D’Cunha et al. conducted a multicenter prospective trial (GALGA 9761) for clinical stage 1 NSCLC, and reported 14% of patients with pathological stage 2 disease, 13.5% with stage 3 disease, and 0.9% with stage 4 disease(29). Decaluwé et al. reported nodal pN1 and pN2 upstaging in 13.3% and 8.2% of their cohort(30). In our study, among 1,735 patients who were preoperative diagnosed with cN0 disease, 171(9.9%) patients had pN1, 191(11.0%) had p(N1+N2), and 91(5.2%) patients had SN2 postoperatively.

Previous studies have identified the following significant associations for nodal upstaging: non-upper located tumors(22), tumor size(31), consolidation with solid component(32), visceral pleural involvement(33), positive tumor markers(34), and tumors with poor differentiation(21,35). Despite this, results from individual studies are not generalizable to other clinics. The large variation in different studies’ models reflects variation in the clinical variables recorded for each patient, variation in criteria for selection of patients, variation in nodal dissection expertise, among other variations. Until pre-specified risk stratification and algorithm are tested in a multi-center prospective trial, this work (and others before it) cannot be used to guide management.

### What does this work add to the field?

Unbalanced datasets are not unique to upstaging models, but the challenges to data analysis have been largely disregarded in prior papers. Using a unique data analysis approach, we improved the characterization of the models, applying 3 equations to model 3 possible outcomes from these patients. Since patients’ selection criteria was specific to cN0 rather than overall stage, this dataset thereby included patients with larger tumor sizes, which allowed us to use tumor size as an independent variable in the model. The variables in the dataset are obtained from low-dose CT imaging and demographic findings, which enable the use of our predictive model in low-resource settings.

### Is there anything that we need to do as clinicians and surgeons for case management?

We hope clinicians will be able to estimate nodal upstaging risk for each clinical nodal negative patient using preoperative parameters, and through this, avoid unnecessary invasive procedure for low risk patients, and also consider more comprehensive nodal dissection for higher risk patients.

The current study has several limitations. The retrospective nature of the dataset limits the number of clinical variables that could be potentially be collected on the patients. Similarly, promising biomarkers could not be tested in a retrospective study. Prospective studies that systematically test the utility of individualized risk assessment for selecting biopsies in cN0 NSCLC patients would be welcome. We welcome collaborations with other centers. Finally, due to the indolent nature of early-stage NSCLC, a long follow-up is needed, and we could not perform a survival analysis at this time.

## Conclusion

In a large retrospective single-institution dataset of 1735 cN0 NSCLC patients, we report an overall 26.1% nodal upstaging rate. 9.9% to pN1, 11.0% to p(N1+N2) and 5.2% to pSN2. After a thorough analysis of multiple clinical factors, we established 3 accurate and easy-to-use predictive models for specific pN upstaging. The predictive models can stratify cN0 NSCLC patients into different risk groups, which can aid clinical management. External validation in a prospective multi-site study is needed before adoption into clinical practice.

## Data Availability

All data and data analysis tools are included in the submission.

## Ethical approval

All procedures performed in studies involving human participants were in accordance with the ethical standards of the institutional and/or national research committee and with the 1964 Helsinki declaration and its later amendments or comparable ethical standards.

## IRB approval

Approval for this retrospective chart review was granted by the Institutional Review Board at the West China Hospital, Sichuan University.

## ABBREVIATION LIST

NSCLC: non-small cell lung cancer
pN: pathological node stage
cN: clinical node stage
CT: computed tomography
PET-CT: positron emission tomography-computed tomography
VATS: video-assisted thoracic surgery
FS: frozen specimen
AUC: area under the curve
RUL: right upper lobe
RML: right middle lobe
RLL: right lower lobe
LUL: left upper lobe
LLL: left lower lobe
GGO: ground-glass opacity
OR: odds ratio
CI: confidence interval
EBUS-TBNA: endobronchial ultrasound-transbronchial needle aspiration
EUB-FNA: endoscopic ultrasound-fine needle aspiration

